# Understanding the cervical cancer self-collection preferences of women living in urban and rural Rwanda

**DOI:** 10.1101/2023.06.15.23291471

**Authors:** Varun Nair, Hallie Dau, Marianne Vidler, Maryam AboMoslim, Barbra Mutamba, McKerron Scott, Zoey Nesbitt, John Deodatha, Schadrack Danson Byiringiro, Charles Niyotwiringiye, Nadia Mithani, Laurie Smith, Gina Ogilvie, Stephen Rulisa

**Author notes:** **Corresponding author:** Gina Ogilvie, MD MSc FCFP DrPH, BC Women’s Hospital and Health Centre, Box 42, Room H203G - 4500 Oak Street, Vancouver, BC, V6H 3N1.

## Abstract

**PURPOSE:** Cervical cancer is a leading cause of cancer among women in low- and middle-income countries. Women in Rwanda have high rates of cervical cancer due to limited access to effective screening methods. Research in other low-resource settings similar to Rwanda has shown that HPV-based self-collection is an effective cervical cancer screening method. This study aims to compare the preferences of Rwandan women in urban and rural settings toward self-collection and to report on factors related to self-collection amenability.

**METHODS:** A cross-sectional survey was conducted from June 1-9, 2022. Women were recruited from one urban and one rural clinic in Rwanda. Women were eligible for the study if they were ≥ 18 years and spoke Kinyarwanda or English. The survey consisted of 51 questions investigating demographics and attitudes towards self-collection for cervical cancer screening. We reported descriptive statistics stratified by urban and rural sites.

**RESULTS:** In total, 169 urban and 205 rural women completed the survey. The majority of respondents at both sites had a primary school or lower education and were in a relationship. Both urban and rural respondents were open to self-collection; however, rates were higher in the rural site (79.9% urban and 95.6% rural; p-value<0.001). Similarly, women in rural areas were more likely to report feeling unembarrassed about self-collection (65.3% of urban, 76.8% of rural; p-value<0.001). Notably, almost all urban and rural respondents (97.6% urban and 98.5% rural) stated they would go for a cervical cancer pelvic examination to a nearby health center if their self-collected results indicated any concern (p-value=0.731).

**CONCLUSION:** Rwandan women in both urban and rural areas largely support self-collection for cervical cancer screening. Further research is needed to better understand how to implement self-collection screening services in Rwanda.

## INTRODUCTION

Cervical cancer is a leading cause of morbidity and mortality in low- and middle-income countries (LMIC), despite being a largely preventable disease. Compared to high-income countries (HIC), LMIC bear a highly disproportionate burden of cervical cancer with 85% of global deaths(1,2). This disproportionate impact in LMICs can, in part, be attributed to limitations in access to appropriate and timely primary prevention, screening, diagnostic, and treatment services(2,3). Due to these limitations, women in LMICs are often diagnosed with an advanced stage of the disease(4).

In 2020, the World Health Organization (WHO) called for the elimination of cervical cancer(5). One of the key goals of this initiative is to have 70% of women screened with a high-performance test by 35 years of age and again by 45 years of age(5). Currently, visual inspection with acetic acid (VIA) is the most common method used to screen women in LMICs(6). However, due to its low sensitivity and need for a healthcare professional to administer the test it poses challenges in resource limited settings (6–8). Human papillomavirus (HPV)-based self-collection has emerged as a reliable alternative in LMIC contexts(9–13). Self-collected vaginal samples for cervical screening allow women to self-screen under the supervision of a healthcare professional or independently(14). HPV-based screening methods are highly effective in the early detection of cervical cancer (15,16). Furthermore, self-collection-based cervical cancer screening methods have shown to be highly acceptable among women in LMICs (13,17–19).

HPV-based self-collection offers the opportunity to improve equitable access to cervical screening by reducing multiple barriers, offering flexibility in programming and being cost-effective in resource poor settings. The WHO in 2022 recommended self-collection for cervical cancer screening to combat cervical cancer in their guidance on self-care interventions for improving health and well-being(20).

Within the context of Eastern Africa, Rawat et al. in 2021 examined the acceptability of and women’s preferences for self-collection in cervical cancer screening in Uganda. They found that self-collection was acceptable if offered with the convenience of users in mind, such as at beneficial times and locations (11). Furthermore, in 2019, Munoru et al. found in Kenya that such integration of self-collection for cervical cancer screening into primary care services empowers the women seeking these services to be champions of their own health by improving their knowledge of cervical cancer, increasing their amenability to being screened, and boosting overall health outcomes(21).

Rwanda has high incidence rates of pre-cancer and invasive cervical cancer (59 per 100,000 and 17 per 100,000, respectively)(22). This can be attributed in part to a lack of wide-spread available screening(22). Niyonsenga et al. reported in their 2021 cross-sectional study of urban Rwandan women an estimated screening access rate of 28.3% in the capital city of Kigali(23). HPV-based self-collection for cervical cancer screening has the potential to help expand access to cervical cancer screening in Rwanda. In order to implement this type of screening program, it is important to understand the preferences of urban and rural women in Rwanda in order to ensure that such a program would benefit all its users. This study sought to compare and describe the preferences of Rwandan women in urban and rural settings towards self-collection-based cervical cancer screening and to report on factors related to self-collection amenability.

## METHODS

### Design, setting, and study population

This was a cross-sectional study using convenience sampling at Muhima and Nyamata District Hospitals in Rwanda, which represent the urban and rural sites, respectively. Women were recruited from June 1-9, 2022 and were eligible if they were 18 years of age and over, spoke English or Kinyarwanda, and were able to provide informed consent. Participants were recruited from the waiting room at each site by four study team members. Surveys were administered in a separate location to maximize privacy of participants and their responses. All participants provided informed consent. Survey administrators were not involved in the direct care of participants and had no prior relationships with participants. Participant responses were captured by study team members on electronic tablets and entered directly into a REDCap(24), a secure web application for research data management. Respondents were compensated 1050 Rwanda Franc ($1 USD) for their involvement in the study.

### Survey

The survey contained 51 questions which included demographics, behaviours, attitudes and knowledge of cervical cancer, cervical cancer screening and follow-up history, knowledge of HPV vaccination, preferences on integrated service delivery for HPV screening, and attitudes toward self-collection at home. Survey questions were informed by and constructed using two survey instruments. The first was the Improving Data for Decision Making in Global Cervical Cancer Programs Toolkit-Part 2 (IDCCP), which is a resource that consolidates operational resources for improving access to high-quality data for decision-making when it comes to constructing cervical cancer programs in LMIC(25). Secondly, a questionnaire administered in Kisenyi, Uganda by Mitchell et al. in 2017 further informed various sections of this study’s survey, such as the behaviours, screening and follow-up, and attitudes towards self-collection at home sections(12). Mitchell et al.’s survey included questions on women’s intentions to provide self-collected samples for cervical cancer screening and were reviewed by local and international experts in women’s health(12), allowing us to seamlessly adapt its items to this survey.

The primary outcome for this analysis is the respondents’ willingness to self-collect for cervical cancer screening. During data collection, research assistants read a script to respondents describing the self-collection procedure. This was done in order to provide context to respondents who were not aware of what self-collection entailed. Responses were recorded on a five-point Likert scale and included: very unlikely, somewhat unlikely, neither likely nor unlikely, somewhat likely, and very likely; or disagree, not sure, and agree. Additional questions on attitudes towards self-collection included willingness to have an outreach worker bring self-collection kits to respondents’ residences and willingness to go to the nearest health center themselves to deposit the swab after self-collection.

Furthermore, women were asked if they would: be embarrassed to perform self-collection at home, be worried they would not be able to perform self-collection correctly, be afraid of receiving a cervical cancer diagnosis after testing themselves, be afraid that other people would think they had HPV or cervical cancer if respondents told them they had tested themselves, be afraid that performing self-collection with a swab would be painful, need their partner’s approval to perform self-collection, and if their religion/spiritual beliefs would affect their decision to be screened for cervical cancer. These were similarly measured using a three-point Likert scale (disagree, not sure, agree, declined to answer). Participants were also asked whether they would go to a clinic for a pelvic examination for cervical cancer if their self-collected sample indicated any concern (response options: I would go, not sure, and I would not go).

Demographic variables captured included clinic location (urban or rural), age, marital status (in a relationship and not in a relationship), education level (less than primary school education and primary school education or above), religious identity (Protestant, Catholic, and other), number of partners, age at first intercourse (less than 18 years and 18 years or above), number of pregnancies, age at first pregnancy, history of chronic diseases (malaria, respiratory disease, HIV/AIDS, high blood pressure, hepatitis, heart disease, tuberculosis, and cancer), and history of health services accessed (child’s health, acute care, antenatal care (ANC), sexual/reproductive health, malaria, HIV/ARV, chronic conditions, and tuberculosis).

### Data analysis

Bivariate descriptive statistics for all survey questions were generated for all respondents by stratifying the respondents by urban and rural sites. Missing data for all questions analyzed in this study were included during analysis and reported in this study’s findings. Fisher’s exact tests were used to compare differences in response between urban and rural respondents. An alpha-value (*α*) of 0.05 was used to determine the significance of inter-site comparisons, with *p <* α indicating a statistically significant difference(26). All statistical analysis was completed using R (version 4.0.3) (27).

### Ethics statement

This study received ethics approvals from the University of British Columbia Research Ethics Board (H22-01335) and the Rwanda National Ethics Committee (125/RNEC/2022). Formal written consent was obtained from all participants. Authors did not have access to information that could identify participants either during or after data collection.

## RESULTS

In total, 374 respondents completed the survey, with 169 respondents from the urban site (Muhima; 45.2%) and 205 respondents from the rural site (Nyamata; 54.8%) (Table 1). The majority of respondents at both sites were between 25-45 years of age (urban n=124, 73.1%; rural n=135, 65.8%; p=0.016), had a primary school or less education (urban n=81, 48.2%; rural n=132, 64.4%, p=0.007), and were in a relationship (urban n=123, 72.8%; rural n=164, 80.4%; p=0.107). Rural respondents were significantly more likely than urban respondents to report having had their first intercourse at or above the age of 18 years (urban n=129, 76.3%; rural n=162, 79.0%; p=0.046). Further details for the demographic characteristics are provided in **Table 1**.

**Table 1.**
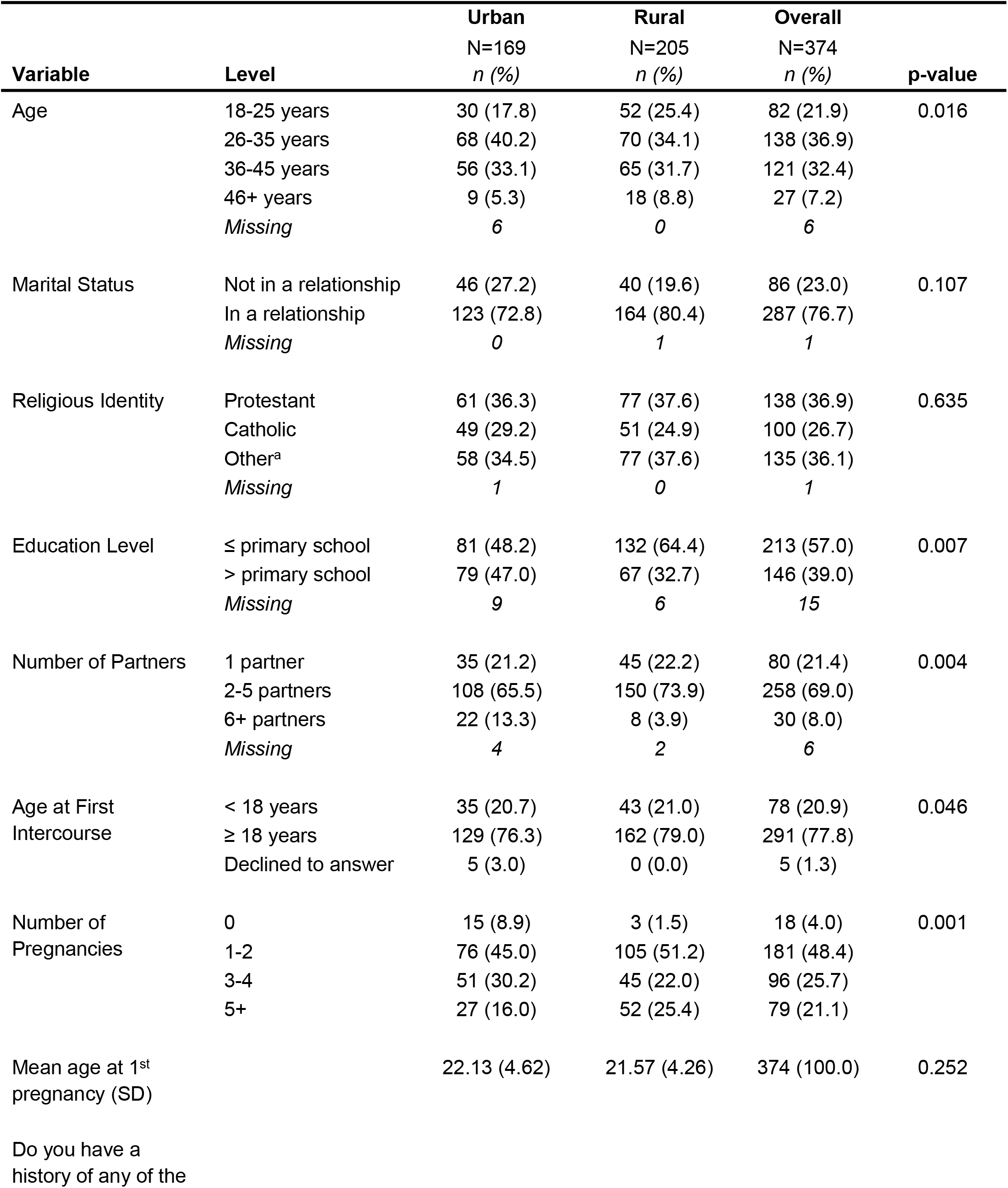
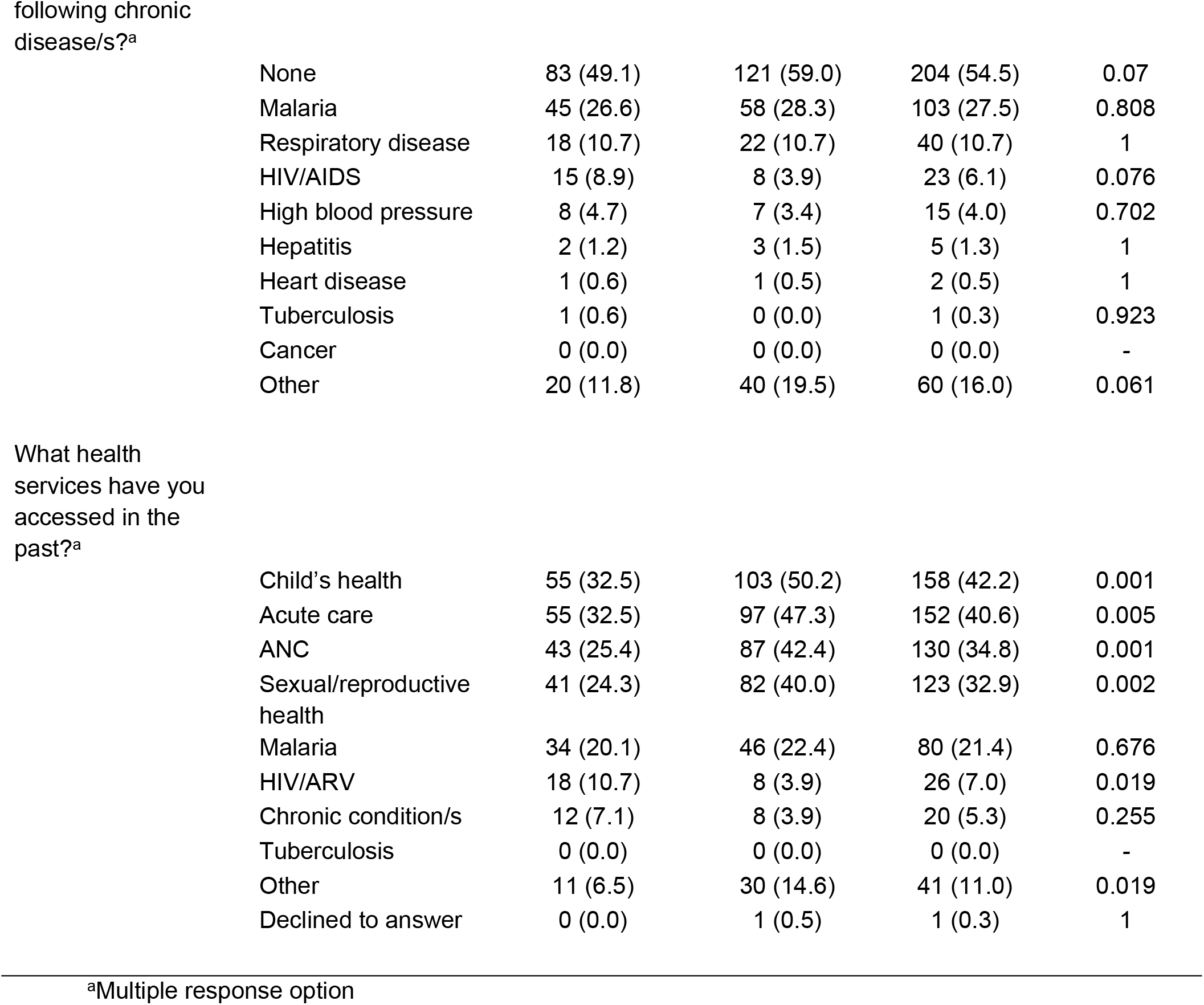
Demographics stratified by urban and rural sites

| Variable | Level | Urban | Rural | Overall | p-value |
| --- | --- | --- | --- | --- | --- |
|  |  | N=169<br>n (%) | N=205<br>n (%) | N=374<br>n (%) |  |
| Age | 18-25 years | 30 (17.8) | 52 (25.4) | 82 (21.9) | 0.016 |
|  | 26-35 years | 68 (40.2) | 70 (34.1) | 138 (36.9) |  |
|  | 36-45 years | 56 (33.1) | 65 (31.7) | 121 (32.4) |  |
|  | 46+ years | 9 (5.3) | 18 (8.8) | 27 (7.2) |  |
|  | Missing | 6 | 0 | 6 |  |
| Marital Status | Not in a relationship | 46 (27.2) | 40 (19.6) | 86 (23.0) | 0.107 |
|  | In a relationship | 123 (72.8) | 164 (80.4) | 287 (76.7) |  |
|  | Missing | 0 | 1 | 1 |  |
| Religious Identity | Protestant | 61 (36.3) | 77 (37.6) | 138 (36.9) | 0.635 |
|  | Catholic | 49 (29.2) | 51 (24.9) | 100 (26.7) |  |
|  | Other<sup>a</sup> | 58 (34.5) | 77 (37.6) | 135 (36.1) |  |
|  | Missing | 1 | 0 | 1 |  |
| Education Level | ≤ primary school | 81 (48.2) | 132 (64.4) | 213 (57.0) | 0.007 |
|  | > primary school | 79 (47.0) | 67 (32.7) | 146 (39.0) |  |
|  | Missing | 9 | 6 | 15 |  |
| Number of Partners | 1 partner | 35 (21.2) | 45 (22.2) | 80 (21.4) | 0.004 |
|  | 2-5 partners | 108 (65.5) | 150 (73.9) | 258 (69.0) |  |
|  | 6+ partners | 22 (13.3) | 8 (3.9) | 30 (8.0) |  |
|  | Missing | 4 | 2 | 6 |  |
| Age at First Intercourse | < 18 years | 35 (20.7) | 43 (21.0) | 78 (20.9) | 0.046 |
|  | ≥ 18 years | 129 (76.3) | 162 (79.0) | 291 (77.8) |  |
|  | Declined to answer | 5 (3.0) | 0 (0.0) | 5 (1.3) |  |
| Number of Pregnancies | 0 | 15 (8.9) | 3 (1.5) | 18 (4.0) | 0.001 |
|  | 1-2 | 76 (45.0) | 105 (51.2) | 181 (48.4) |  |
|  | 3-4 | 51 (30.2) | 45 (22.0) | 96 (25.7) |  |
|  | 5+ | 27 (16.0) | 52 (25.4) | 79 (21.1) |  |
| Mean age at 1<sup>st</sup> pregnancy (SD) |  | 22.13 (4.62) | 21.57 (4.26) | 374 (100.0) | 0.252 |
| Do you have a history of any of the |  |  |  |  |  |

|  |  |  |  |  |
| --- | --- | --- | --- | --- |
| None | 83 (49.1) | 121 (59.0) | 204 (54.5) | 0.07 |
| Malaria | 45 (26.6) | 58 (28.3) | 103 (27.5) | 0.808 |
| Respiratory disease | 18 (10.7) | 22 (10.7) | 40 (10.7) | 1 |
| HIV/AIDS | 15 (8.9) | 8 (3.9) | 23 (6.1) | 0.076 |
| High blood pressure | 8 (4.7) | 7 (3.4) | 15 (4.0) | 0.702 |
| Hepatitis | 2 (1.2) | 3 (1.5) | 5 (1.3) | 1 |
| Heart disease | 1 (0.6) | 1 (0.5) | 2 (0.5) | 1 |
| Tuberculosis | 1 (0.6) | 0 (0.0) | 1 (0.3) | 0.923 |
| Cancer | 0 (0.0) | 0 (0.0) | 0 (0.0) | - |
| Other | 20 (11.8) | 40 (19.5) | 60 (16.0) | 0.061 |

|  |  |  |  |  |
| --- | --- | --- | --- | --- |
| Child's health | 55 (32.5) | 103 (50.2) | 158 (42.2) | 0.001 |
| Acute care | 55 (32.5) | 97 (47.3) | 152 (40.6) | 0.005 |
| ANC | 43 (25.4) | 87 (42.4) | 130 (34.8) | 0.001 |
| Sexual/reproductive<br>health | 41 (24.3) | 82 (40.0) | 123 (32.9) | 0.002 |
| Malaria | 34 (20.1) | 46 (22.4) | 80 (21.4) | 0.676 |
| HIV/ARV | 18 (10.7) | 8 (3.9) | 26 (7.0) | 0.019 |
| Chronic condition/s | 12 (7.1) | 8 (3.9) | 20 (5.3) | 0.255 |
| Tuberculosis | 0 (0.0) | 0 (0.0) | 0 (0.0) | - |
| Other | 11 (6.5) | 30 (14.6) | 41 (11.0) | 0.019 |
| Declined to answer | 0 (0.0) | 1 (0.5) | 1 (0.3) | 1 |

When asked if they would be willing to independently self-collect samples for cervical cancer screening at home if they were provided with the appropriate instruction, 135 (79.9%) urban and 196 (95.6%) rural respondents expressed either very or somewhat willing (p<0.001) **(Table 2)**. Similarly, 139 (82.2%) urban respondents and 193 (94.6%) rural respondents were in favor of having a community outreach worker supply them with a self-collection swab at their home (p<0.001). When asked about their willingness to drop off their self-collected sample at the health centre closest to them, 134 (79.3%) urban respondents and 192 (93.7%) rural respondents expressed a willingness to do so (p<0.001). Additionally,156 (76.8%) rural respondents disagreed with the notion that they would be embarrassed to perform self-collection procedures at home (p<0.001). Furthermore, 139 (68.1%) rural respondents disagreed when presented the statement that they would be afraid that self-collection would show they had cervical cancer (p=0.039). Interestingly, 96 (56.8%) rural respondents and 142 (69.6%) urban respondents disagreed that they were worried about incorrectly performing the self-collection procedure (p<0.001). In the most overwhelmingly positive response regarding self-collection for cervical cancer screening in this survey, 162 (97.6%) urban respondents and 200 (98.5%) rural respondents expressed willingness to go for a pelvic examination to check for cervical cancer following self-collection if, upon further analysis, their self-collected sample indicated any concern (p=0.731).

**Table 2.**
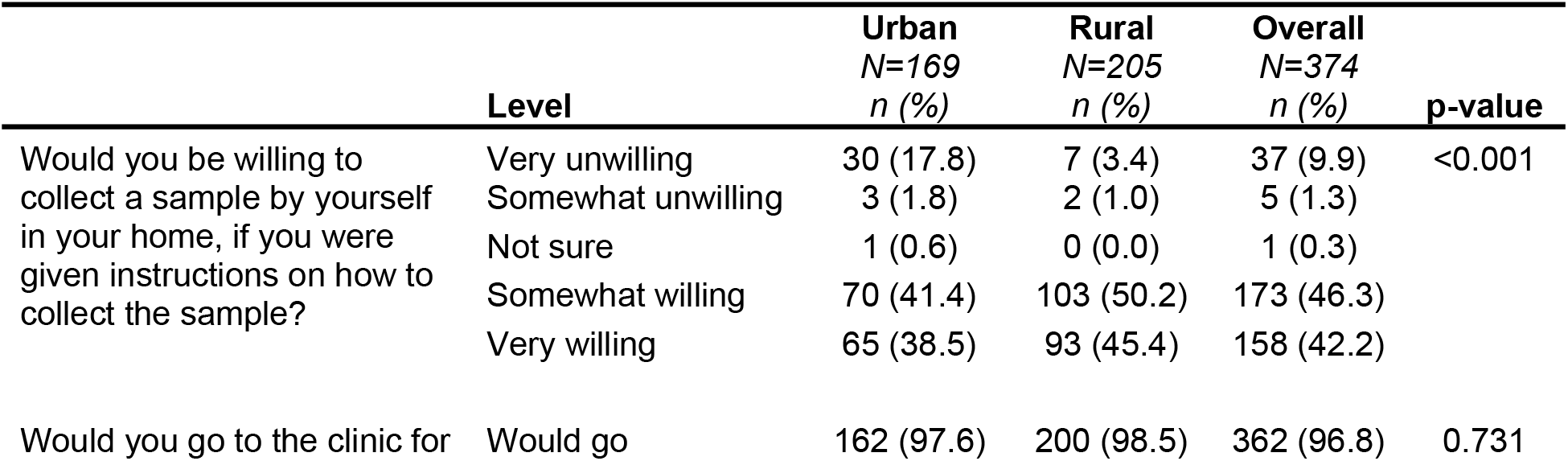
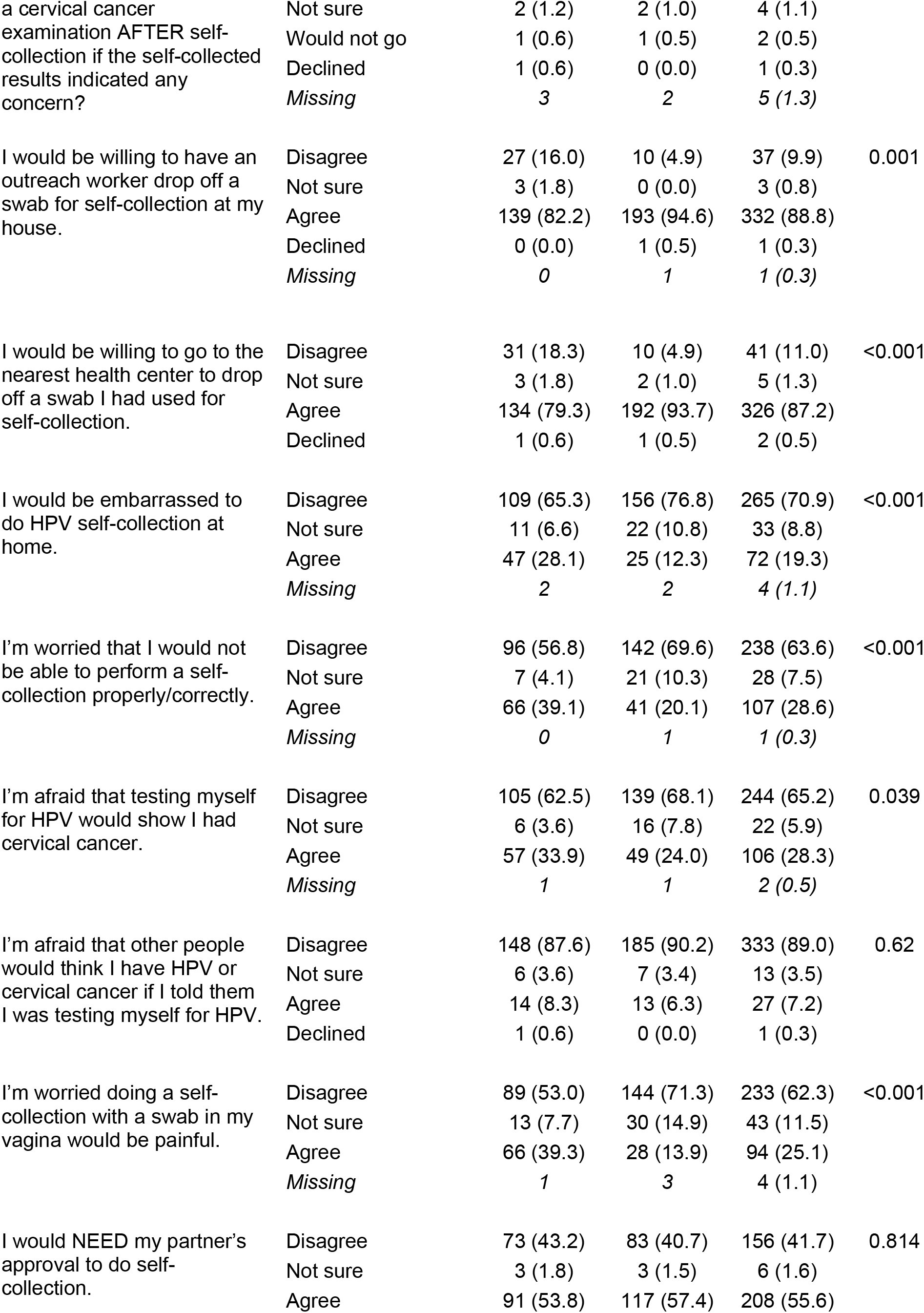
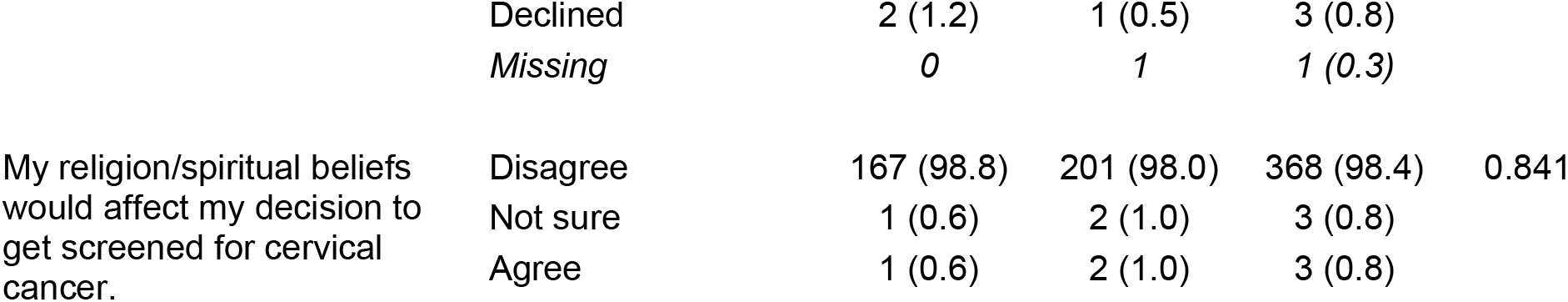
Responses to questions investigating willingness to self-collect cervical samples stratified by urban and rural sites

## DISCUSSION

In this study, we evaluated willingness to self-collect for cervical screening from women in Muhima and Nyamata District Hospitals. We found that women in both urban and rural settings were willing to self-collect for cervical cancer screening. Furthermore, women reported that they would be able to perform the test given proper instruction with women in rural areas being statistically more willing than urban.

We found that the majority of women in both urban and rural Rwanda were willing to self-collect at home for cervical cancer screening. This finding aligns with current research on self-collection in low-resource settings(12),(28),(29). For example, a 2011 cross-sectional study in Kisenyi, Uganda by Mitchell et al. reported that 80.6% of Ugandan women were willing to self-collect for cervical cancer screening(12). Our findings in rural Rwanda also aligned with similar research conducted in other rural, low-resource settings, such a studies conducted by Swanson et al. in rural Kenya(30), Gottschlich et al. in rural Guatemala(31), and Mremi et al. in rural Tanzania(32). That women at both urban and rural areas exhibited such positivity towards self-collection is highly encouraging for the prospects of implementing formal self-collection-based cervical cancer screening program which in turn could improve access to consistent cervical cancer screening for women in Rwanda. Our finding that rural respondents are significantly more willing to participate in self-collection warrants further research to understand why there might exist a difference between urban and rural settings in Rwanda. This finding also tells us that a uniformly-delivered self-collection-based cervical cancer screening program in Rwanda would have different levels of uptake in urban and rural areas, prompting the need to for the program to be built such that it is contextually adapted to barriers in different regions.

Women in both of urban and rural areas were willing to have a community outreach worker drop off a self-collection kit at their homes and were similarly willing to go to a nearby health centre to deposit their self-collected samples. From this, it is evident that the logistics of self-collection do not negatively impact Rwandan women’s amenability to self-collect, nor do they impact their willingness to continue along the care pathway. This latter finding is similar to the case in rural Uganda, where Rawat et al. found that 83% of women were willing to have a community health worker (CHW) drop off self-collection kits at their homes(11). Arrossi et al. also found CHW-led drop-off of self-collection resources to be both acceptable in their evaluation of a self-collection-based cervical cancer screening program in rural Argentina(33). Interestingly, our findings in urban Rwanda differ from research conducted by Ogilvie et al. in urban Uganda, where 68.7% of respondents reported an unwillingness to go to a nearby health centre to deposit their self-collected samples(34). Little research exists on the acceptability of CHW-led drop-off of self-collection resources in urban LMIC settings, making this finding an area for further research. Even so, given the positivity of our research in this regard in both urban and rural Rwanda, our results are highly encouraging in showcasing the potential success of an integrated self-collection-based cervical cancer screening program for women in Rwanda wherein CHWs are the leaders of resource delivery to enable self-collection to occur.

Our findings demonstrate that social stigma and religious beliefs would not deter women from self-collecting. However, we did find that the majority of participants at both sites believed that they would need their partner’s approval to self-collect for cervical cancer screening. This presents a notable barrier when considering the implementation of self-collection to cervical cancer screening in Rwanda. In rural Malawi, Lee et al. found that male partners were indeed reported to be a barrier to consistently accessing a self-collection-based cervical cancer screening program by women in their study(35). Recent studies by Moucheraud et al. in Malawi in 2020(36), Binka et al. in Ghana in 2019(37), and Buchanan Lunsford et al.(38) and Adewumi et al.(39) in Kenya in 2017 and 2019, respectively, also corroborate this notion that male partner approval is an important factor that must be considered and navigated with intention when designing a successful self-collection-based cervical cancer screening program. A successful screening program in Rwanda may not be possible without support from partners. As such, continued exploration must seek to understand the views of partners regarding self-collection in Rwanda, and what steps are necessary to ensure greater access to a self-collection-based cervical cancer screening program in Rwanda.

This study is strengthened by the expertise of the team conducting the project. The study team is highly knowledgeable in cervical cancer self-collection and is familiar working in low-resource settings. Furthermore, local research assistants collected the data in respondents’ native languages and were familiar with the region and data collection procedures. The use of convenience sampling remains a limitation as it can contribute to sampling bias(40).

Furthermore, as the data was self-reported, it is possible that there was some misreporting either due to social desirability or recall bias from the respondents(41). Lastly, given that our respondents were recruited from only two sites overall and only one site each from urban and rural settings, our study population may not be representative of the true population of Rwanda, thereby limiting the broader applicability of our results across Rwanda.

In conclusion, the results from our study show that women in Rwanda show a clear support for self-collection as a means of sampling for cervical cancer screening. As such, further research is needed on how to most effectively implement a such a program in Rwanda. An accessible cervical cancer screening program that is amenable for all Rwandan women is crucial to not only eliminate cervical cancer in Rwanda, but worldwide.

## Data Availability

Data is not available for access as it contains human data that contains potentially identifying information about participants. All data inquires can be made to Gina Ogilvie

## Acknowledgements

We would like to thank Muhima and Nyamata District Hospitals for their assistance in this study.

## References

1. Cervical cancer [Internet]. World Health Organization; 2022 [cited 2022 Nov 13]. Available from: https://www.who.int/news-room/fact-sheets/detail/human-papillomavirus-(hpv)-and-cervical-cancer

2. Parkin DM, Bray F, Ferlay J, Pisani P. Global Cancer Statistics, 2002. CA Cancer J Clin. 2005 Mar 1;55(2):74–108.

3. Katahoire RA, Jitta J, Kivumbi G, Murokora D, Arube WJ, Siu G, et al. An assessment of the readiness for introduction of the HPV vaccine in Uganda. Afr J Reprod Health. 2008 Dec;12(3):159–72.

4. Vu M, Yu J, Awolude OA, Chuang L. Cervical cancer worldwide. Curr Probl Cancer. 2018 Sep;42(5):457–65.

5. WHO Global strategy to accelerate the elimination of cervical cancer as a public health problem [Internet]. World Health Organization; 2020 [cited 2023 Mar 4]. Available from: https://www.who.int/publications/i/item/9789240014107

6. Hull R, Mbele M, Makhafola T, Hicks C, Wang S, Reis R, et al. Cervical cancer in low and middle-income countries (Review). Oncol Lett. 2020 Jun 19;20(3):2058–74.

7. Senapathy JG, Umadevi P, Kannika PS. The present scenario of cervical cancer control and HPV epidemiology in India: an outline. Asian Pac J Cancer Prev APJCP. 2011;12(5):1107–15.

8. Mishra GA, Pimple SA, Shastri SS. Prevention of Cervix Cancer in India. Oncology. 2016;91(Suppl. 1):1–7.

9. Ndayisaba G, Verwijs MC, van Eeckhoudt S, Gasarabwe A, Hardy L, Borgdorff H, et al. Feasibility and Acceptability of a Novel Cervicovaginal Lavage Self-Sampling Device Among Women in Kigali, Rwanda. Sex Transm Dis. 2013 Jul;40(7):552–5.

10. Verwijs MC, Agaba S, Umulisa MM, van de Wijgert JHHM. Feasibility and acceptability of frequent vaginal self-sampling at home by Rwandan women at high risk of urogenital tract infections. Sex Transm Infect. 2022 Feb;98(1):58–61.

11. Rawat A, Sanders C, Mithani N, Amuge C, Pedersen H, Namugosa R, et al. Acceptability and preferences for self‐collected screening for cervical cancer within health systems in rural Uganda: A mixed‐methods approach. Int J Gynecol Obstet. 2021 Jan;152(1):103–11.

12. Mitchell SM, Pedersen HN, Eng Stime E, Sekikubo M, Moses E, Mwesigwa D, et al. Self-collection based HPV testing for cervical cancer screening among women living with HIV in Uganda: a descriptive analysis of knowledge, intentions to screen and factors associated with HPV positivity. BMC Womens Health. 2017 Dec;17(1):4.

13. Mitchell S, Ogilvie G, Steinberg M, Sekikubo M, Biryabarema C, Money D. Assessing women’s willingness to collect their own cervical samples for HPV testing as part of the ASPIRE cervical cancer screening project in Uganda. Int J Gynecol Obstet. 2011 Aug;114(2):111–5.

14. Lofters A, Vahabi M. Self-sampling for HPV to enhance uptake of cervical cancer screening: Has the time come in Canada? Can Med Assoc J. 2016 Sep 6;188(12):853–4.

15. Nishimura H, Yeh PT, Oguntade H, Kennedy CE, Narasimhan M. HPV self-sampling for cervical cancer screening: a systematic review of values and preferences. BMJ Glob Health. 2021 May;6(5):e003743.

16. Nakisige C, Trawin J, Mitchell-Foster S, Payne BA, Rawat A, Mithani N, et al. Integrated cervical cancer screening in Mayuge District Uganda (ASPIRE Mayuge): a pragmatic sequential cluster randomized trial protocol. BMC Public Health. 2020 Dec;20(1):142.

17. Esber A. Feasibility, validity and acceptability of self-collected samples for human papillomavirus (HPV) testing in rural Malawi. Malawi Med J. 2018 Jun 30;30(2):61.

18. Broquet C, Triboullier D, Untiet S, Schafer S, Petignat P, Vassilakos P. Acceptability of self-collected vaginal samples for HPV testing in an urban and rural population of Madagascar. Afr Health Sci. 2015 Sep 9;15(3):755.

19. Murchland AR, Gottschlich A, Bevilacqua K, Pineda A, Sandoval-Ramírez BA, Alvarez CS, et al. HPV self-sampling acceptability in rural and indigenous communities in Guatemala: a cross-sectional study. BMJ Open. 2019 Oct;9(10):e029158.

20. Recommendations and key considerations [Internet]. WHO guideline on self-care interventions for health and well-being, 2022 revision. World Health Organization; 2022 [cited 2023 Apr 8]. Available from: https://www.ncbi.nlm.nih.gov/books/NBK582350/

21. Munoru F, Gitonga L, Muraya M. Integration of Cervical Cancer Screening Services in the Routine Examinations Offered in the Kenyan Health Facilities: A Systematic Review. Open J Obstet Gynecol. 2019;09(05):656–68.

22. Makuza JD, Nsanzimana S, Muhimpundu MA, Pace LE, Ntaganira J, Riedel DJ. Prevalence and risk factors for cervical cancer and pre-cancerous lesions in Rwanda. Pan Afr Med J [Internet]. 2015 [cited 2023 Mar 7];22. Available from: http://www.panafrican-med-journal.com/content/article/22/26/full/

23. Niyonsenga G, Gishoma D, Sego R, Uwayezu MG, Nikuze B, Fitch M, et al. Knowledge, utilization and barriers of cervical cancer screening among women attending selected district hospitals in Kigali - Rwanda. Can Oncol Nurs J. 2021 Jul 22;31(3):266–74.

24. Patridge EF, Bardyn TP. Research Electronic Data Capture (REDCap). J Med Libr Assoc [Internet]. 2018 Jan 12 [cited 2023 Mar 7];106(1). Available from: http://jmla.pitt.edu/ojs/jmla/article/view/319

25. World Health Organization. Improving data for decision-making: a toolkit for cervical cancer prevention and control programmes [Internet]. Geneva: World Health Organization; 2018 [cited 2023 Mar 7]. 289 p. Available from: https://apps.who.int/iris/handle/10665/279420

26. Infanger D, Schmidt‐Trucksäss A. P value functions: An underused method to present research results and to promote quantitative reasoning. Stat Med. 2019 Sep 20;38(21):4189–97.

27. R Core Team. R: A Language and Environment for Statistical Computing [Internet]. Vienna, Austria; 2021. Available from: https://www.R-project.org/

28. Bansil P, Wittet S, Lim JL, Winkler JL, Paul P, Jeronimo J. Acceptability of self-collection sampling for HPV-DNA testing in low-resource settings: a mixed methods approach. BMC Public Health. 2014 Dec;14(1):596.

29. Gottschlich A, Nuntadusit T, Zarins KR, Hada M, Chooson N, Bilheem S, et al. Barriers to cervical cancer screening and acceptability of HPV self-testing: a cross-sectional comparison between ethnic groups in Southern Thailand. BMJ Open. 2019 Nov;9(11):e031957.

30. Swanson M, Ibrahim S, Blat C, Oketch S, Olwanda E, Maloba M, et al. Evaluating a community-based cervical cancer screening strategy in Western Kenya: a descriptive study. BMC Womens Health. 2018 Dec;18(1):116.

31. Gottschlich A, Rivera-Andrade A, Grajeda E, Alvarez C, Mendoza Montano C, Meza R. Acceptability of Human Papillomavirus Self-Sampling for Cervical Cancer Screening in an Indigenous Community in Guatemala. J Glob Oncol. 2017 Oct;3(5):444–54.

32. Mremi A, Linde DS, Mchome B, Mlay J, Schledermann D, Blaakær J, et al. Acceptability and feasibility of self‐sampling and follow‐up attendance after text message delivery of human papillomavirus results: A cross‐sectional study nested in a cohort in rural Tanzania. Acta Obstet Gynecol Scand. 2021 Apr;100(4):802–10.

33. Arrossi S, Paolino M, Thouyaret L, Laudi R, Campanera A. Evaluation of scaling-up of HPV self-collection offered by community health workers at home visits to increase screening among socially vulnerable under-screened women in Jujuy Province, Argentina. Implement Sci. 2017 Dec;12(1):17.

34. Ogilvie GS, Mitchell S, Sekikubo M, Biryabarema C, Byamugisha J, Jeronimo J, et al. Results of a community-based cervical cancer screening pilot project using human papillomavirus self-sampling in Kampala, Uganda. Int J Gynecol Obstet. 2013 Aug;122(2):118–23.

35. Lee F, Bula A, Chapola J, Mapanje C, Phiri B, Kamtuwange N, et al. Women’s experiences in a community-based screen-and-treat cervical cancer prevention program in rural Malawi: a qualitative study. BMC Cancer. 2021 Dec;21(1):428.

36. Moucheraud C, Kawale P, Kafwafwa S, Bastani R, Hoffman RM. “It is big because it’s ruining the lives of many people in Malawi”: Women’s attitudes and beliefs about cervical cancer. Prev Med Rep. 2020 Jun;18:101093.

37. Binka C, Doku DT, Nyarko SH, Awusabo-Asare K. Male support for cervical cancer screening and treatment in rural Ghana. Withers MH, editor. PLOS ONE. 2019 Nov 18;14(11):e0224692.

38. Buchanan Lunsford N, Ragan K, Lee Smith J, Saraiya M, Aketch M. Environmental and Psychosocial Barriers to and Benefits of Cervical Cancer Screening in Kenya. The Oncologist. 2017 Feb 1;22(2):173–81.

39. Adewumi K, Oketch SY, Choi Y, Huchko MJ. Female perspectives on male involvement in a human-papillomavirus-based cervical cancer-screening program in western Kenya. BMC Womens Health. 2019 Dec;19(1):107.

40. Emerson RW. Convenience Sampling, Random Sampling, and Snowball Sampling: How Does Sampling Affect the Validity of Research? J Vis Impair Blind. 2015 Mar;109(2):164–8.

41. Rosenman R, Tennekoon V, Hill LG. Measuring bias in self-reported data. Int J Behav Healthc Res. 2011;2(4):320.

